# Implementing Home-Based Digital Health in Rural Canada: A Scoping Review

**DOI:** 10.1101/2025.08.27.25334390

**Authors:** Joy Lai, Amareena Saleh-Singh, Gladys Olisaekee

**Affiliations:** Institute of Biomedical Engineering, Faculty of Applied Science and Engineering, University of Toronto, Toronto, Ontario, Canada; Munk School of Global Affairs & Public Policy, University of Toronto, Toronto, Ontario, Canada; Dalla Lana School of Public Health, University of Toronto, Toronto, Ontario, Canada

**Keywords:** Home-based digital health, rural health services, Canada, digital health implementation, remote monitoring, community readiness, digital literacy, health equity

## Abstract

**Objective:** This scoping review maps the current evidence on implementing home-based digital health technologies in rural Canada. It examines available readiness tools and indicators, stakeholder perspectives, barriers, and outcomes to provide evidence-based insights for successful implementation.

**Methods:** A comprehensive search was conducted in Ovid MEDLINE, IEEE Xplore, and Scopus between February and March 2025. Eligible studies focused on patient-facing, home-based digital health technologies in rural or remote Canadian contexts. Articles addressing pre-implementation, implementation, or adoption of home-based digital health solutions were also included. Data extraction and thematic analysis were performed to synthesize findings.

**Results:** Sixteen studies met the inclusion criteria, spanning diverse rural regions of British Columbia, Ontario, and several Prairie and Atlantic provinces. Findings were categorized under four major themes: (1) readiness tools, frameworks, and indicators; (2) patient and provider perspectives; (3) barriers and corresponding strategies; and (4) outcomes and impacts of home-based digital health implementation in rural Canada. While patients and providers are generally positive towards home-based digital health technologies, several context-dependent factors influence their success. Key barriers include digital divides, infrastructure limitations, and varying digital literacy. Effective implementation necessitates addressing these challenges through tailored strategies, such as culturally sensitive design, infrastructure development, digital literacy training, and community engagement.

**Conclusion:** Although home-based digital health technologies have the potential to improve healthcare access and outcomes in rural Canada. Successful implementation requires careful consideration of contextual factors, proactive barrier mitigation, and a focus on co-design with users to ensure equitable access and outcomes.

## 1 Introduction

Rural and remote communities in Canada experience persistent health inequities resulting from geographic isolation, limited availability of healthcare professionals, and reduced access to specialist care and diagnostic services (1,2). Although approximately 18 percent of Canadians live in rural or remote areas, these regions remain underserved by healthcare infrastructure, including primary care, mental health services, and chronic disease management programs (1). Barriers such as travel time, cost, weather conditions, and lack of reliable transportation often lead to delayed or missed care, contributing to poorer health outcomes for rural residents (1,3,4).

Digital health technologies, including telehealth, remote patient monitoring, virtual consultations, and mobile health applications, have emerged as promising tools to bridge access gaps in rural settings (5). Increasingly, these tools are being designed for in-home use, enabling patients to connect with healthcare providers, monitor chronic conditions, and receive care remotely (6,7). The COVID-19 pandemic accelerated the adoption of virtual care across various health domains, including chronic disease management, mental health, and palliative care, demonstrating its feasibility and value (8).

However, moving from reactive pandemic-driven solutions to sustainable digital health models requires deliberate planning, comprehensive evaluation, and a strong foundation of community readiness. Implementing digital health technologies in rural Canada presents significant challenges, including persistent digital divides related to infrastructure, affordability, and digital literacy. There are also issues of mistrust in technology or healthcare systems; and cultural or linguistic mismatches, particularly within Indigenous communities (7–9). Additionally, healthcare providers frequently reported frustrations with technology leading to emotional exhaustion, and a lack of alignment between digital tools and existing systems (10,11). While digital health has the potential to promote equity, it also risks reinforcing existing disparities without context-sensitive planning and design.

The existing literature on digital health in rural Canada is fragmented, spanning multiple disciplines. Few studies focus specifically on home-based patient-facing digital health technologies. Healthcare providers and digital health developers are seeking practical, evidence-based insights to guide the implementation of these technologies. They need evidence that identifies where and when these technologies are most likely to succeed, how to effectively prepare communities and providers for digital care, and what outcomes can be expected. Understanding this evidence is essential for developing equitable and scalable digital health solutions in underserved areas.

With this scoping review, we aim to map the available evidence on readiness tools and indicators, patient and provider perspectives, barriers and implementation strategies, and observed outcomes and impacts of home-based digital health technologies in rural Canadian contexts.

### 1.1 Objectives and Review Questions

To map the current state of the literature regarding the implementation of home-based digital health technologies in rural Canada, our review was guided by the following research questions:

- What readiness tools, frameworks, or indicators exist to assess whether rural communities are prepared for home-based digital health technology implementation?
- What are patient and provider attitudes toward these technologies, and how do these attitudes influence adoption and engagement?
- What are the key barriers to implementation, and what strategies have been employed to address them?
- What outcomes and impacts have been documented following the implementation of home-based digital health technologies in rural Canadian communities?

## 2 Methods

This scoping review was guided by Arksey and O’Malley’s (12) five-stage framework, incorporating methodological refinements proposed by Levac et al. The stages include: (1) identifying the research question; (2) identifying relevant studies; (3) selecting studies for data extraction; (4) extracting and charting the data; and (5) summarizing and reporting the findings (13). We also incorporated key elements of the **P**referred **R**eporting **I**tems for **S**ystematic Reviews and **M**eta-**A**nalyses (PRISMA-ScR) guidelines to ensure transparency and rigor in reporting (Moher et al., 2009).

### 2.1 Search Strategy and Data Sources

A comprehensive literature search was conducted across three major databases: Ovid MEDLINE, IEEE Xplore, and Scopus. The search aimed to identify studies relevant to digital health tools designed for use in rural Canadian healthcare settings. Our focus was specifically on patient-facing technologies that can be accessed and used independently from patients’ homes, without the need to travel to clinics or community centers. The search strategies were developed in consultation with an experienced Liaison & Education Librarian (Eden Kinzel, Gerstein Science Information Centre, University of Toronto). Boolean operators were used to combine key concepts related to social, cultural, infrastructural, technological, and equity-related factors (for example “digital divide,” “barriers,” “readiness,” “telehealth”) with geographic identifiers (e.g., “rural,” “remote,” “Indigenous,”) and digital health–specific terms (e.g., “telemedicine,” “eHealth,” “virtual care,” “AI,” “wearable health technology”). These were further linked with geographic terms identifying Canadian populations and regions, including provinces, territories, and major cities. Medical Subject Headings (MeSH) terms were used where appropriate to enhance sensitivity and relevance. All searches were conducted between February and March 2025.

### 2.2 Inclusion and Exclusion Criteria

Eligible studies were peer-reviewed articles published within the past ten years (2015 to 2025) to ensure relevance to contemporary practices and technological advancements (15). The focus was on home-based, patient-facing, digital health interventions that address challenges in rural or remote settings. Digital health interventions are defined as technologies that patients can access and use independently from their homes, such as mobile applications, remote monitoring devices, or home-based telehealth platforms. Only studies conducted in Canada or those focused on Canadian populations were included for contextual relevance. Full-text availability was required for comprehensive data extraction, and only primary research articles, systematic reviews, or meta-analyses were considered. Studies had to describe implementation, patient or provider experiences, readiness assessments, barriers, or outcomes. While the primary inclusion window was 2015 to 2025, earlier studies were included if they directly addressed most of our research questions and provided unique insights relevant to rural digital health implementation. Grey literature, non-English publications, editorials, or commentaries, and studies focused on technologies used solely by professionals (e.g., electronic health records) were excluded.

### 2.3 Study Selection Process

A total of 3,241 records were imported into Covidence: Ovid MEDLINE (n = 2,642), IEEE Xplore (n = 432), and Scopus (n = 167). One additional article was identified through citation searching. After removing 243 duplicates, two reviewers (JL and AS) independently screened 2,998 records by title and abstract. This was followed by a full-text screening of 337 articles conducted by two reviewers, JL and GOO. Conflicts were resolved through discussion, and if consensus could not be reached, a third reviewer acted as an adjudicator.

After full-text screening, 321 articles were excluded. Sixteen studies met all eligibility criteria and were included in the final review. A summary of the study selection is illustrated in the PRISMA flow diagram (Figure 1).

**Figure 1:**
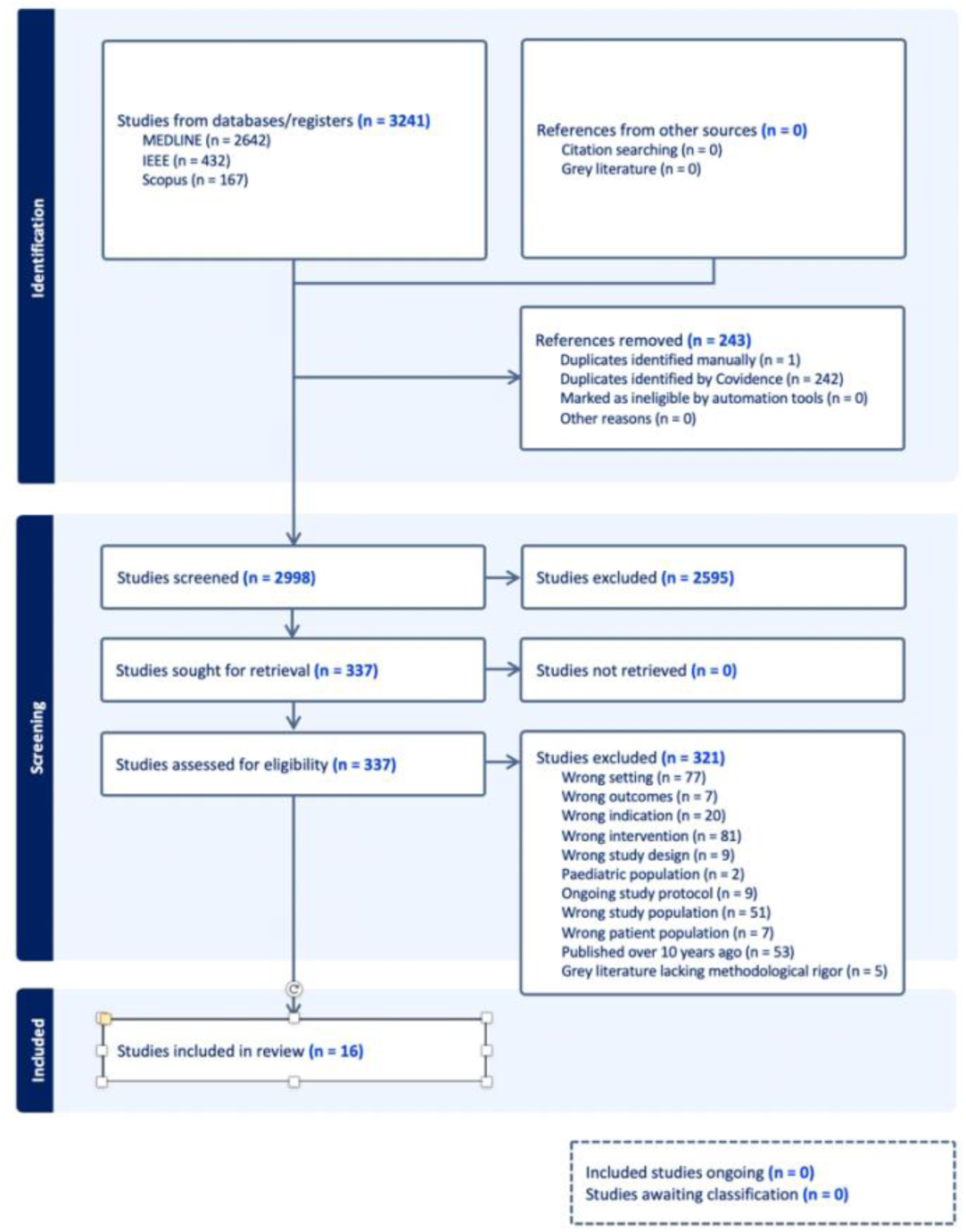
PRISMA flowchart of included studies

### 2.4 Data Extraction and Thematic Analysis

Data from the studies included were extracted using a standardized form in Covidence and organized chronologically in a Microsoft Excel spreadsheet (see Table 1). The key characteristics obtained from each article include the title, authors, publication year, setting, demographic information of the study population, and the review’s primary objectives. Additionally, we noted the main findings and their relevance to in-home digital health services in rural Canada. We conducted a deductive thematic analysis based on our four guiding research questions. The research tream refined themes through ongoing discussions. As a result of this thematic analysis, we categorized the studies into four overarching themes that align with our research questions: (1) Readiness Tools, Frameworks, and Indicators; (2) Patient and Provider Perspectives; (3) Barriers and Corresponding Strategies; and (4) Outcomes and Impacts.

**Table 1:**
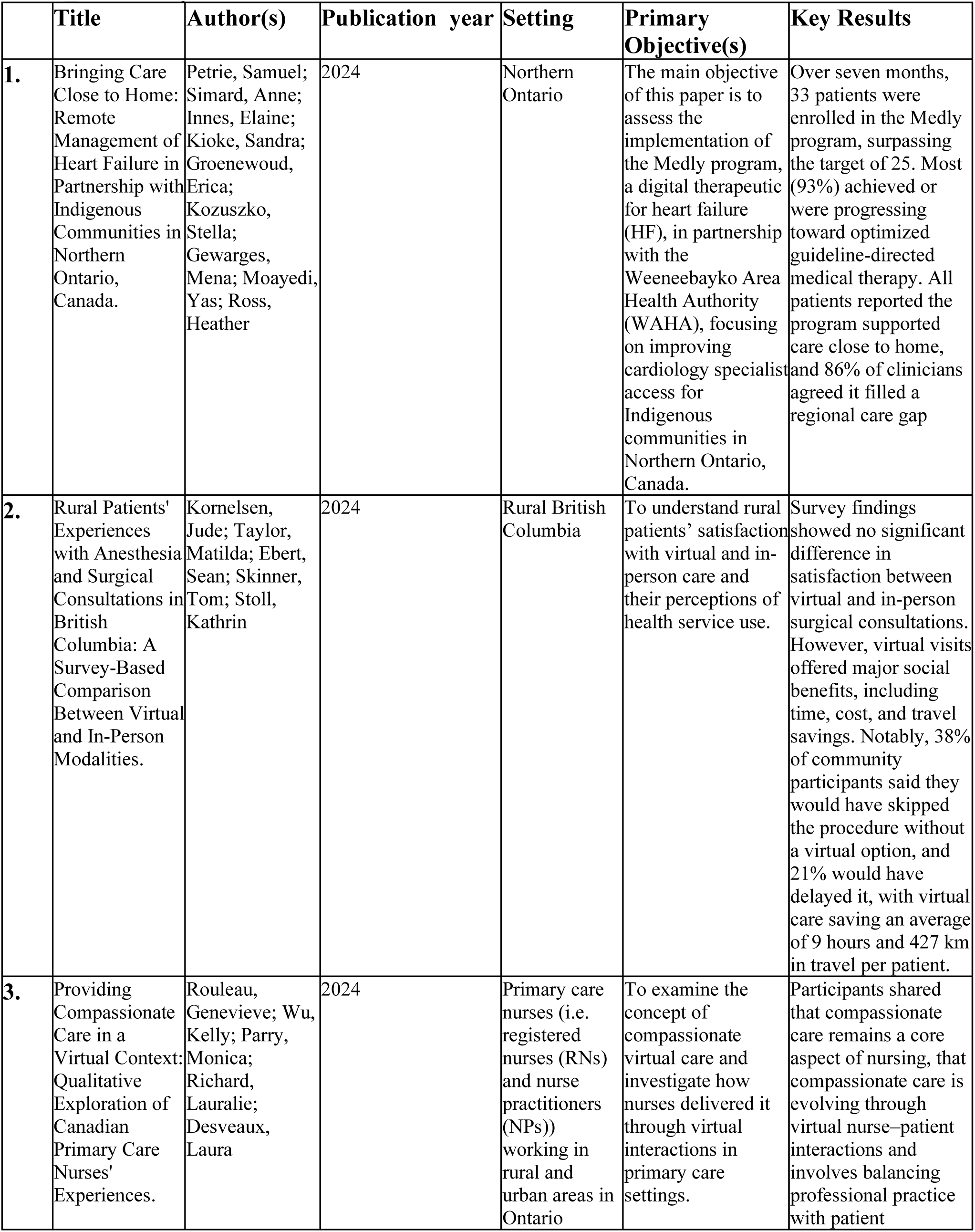
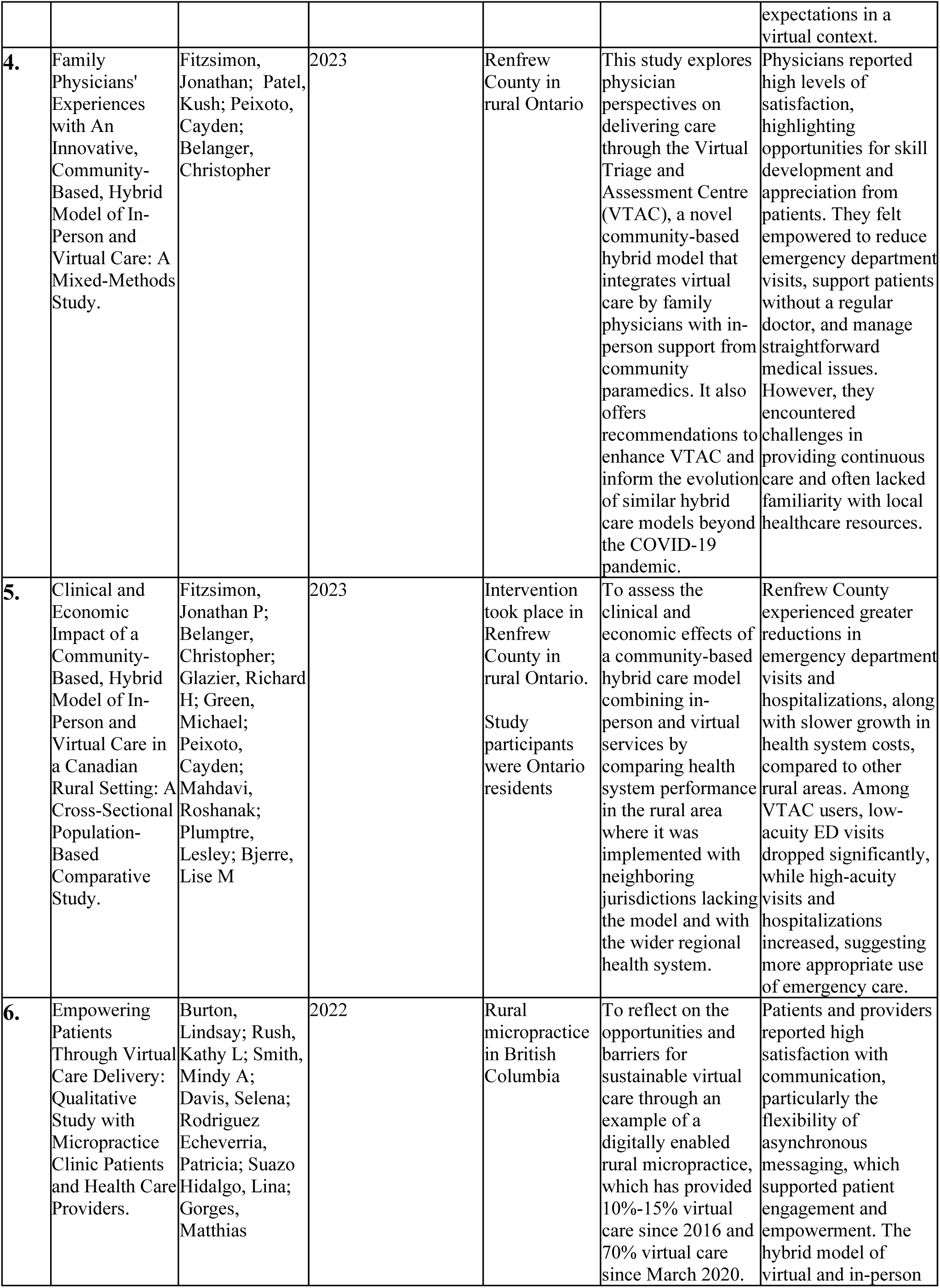
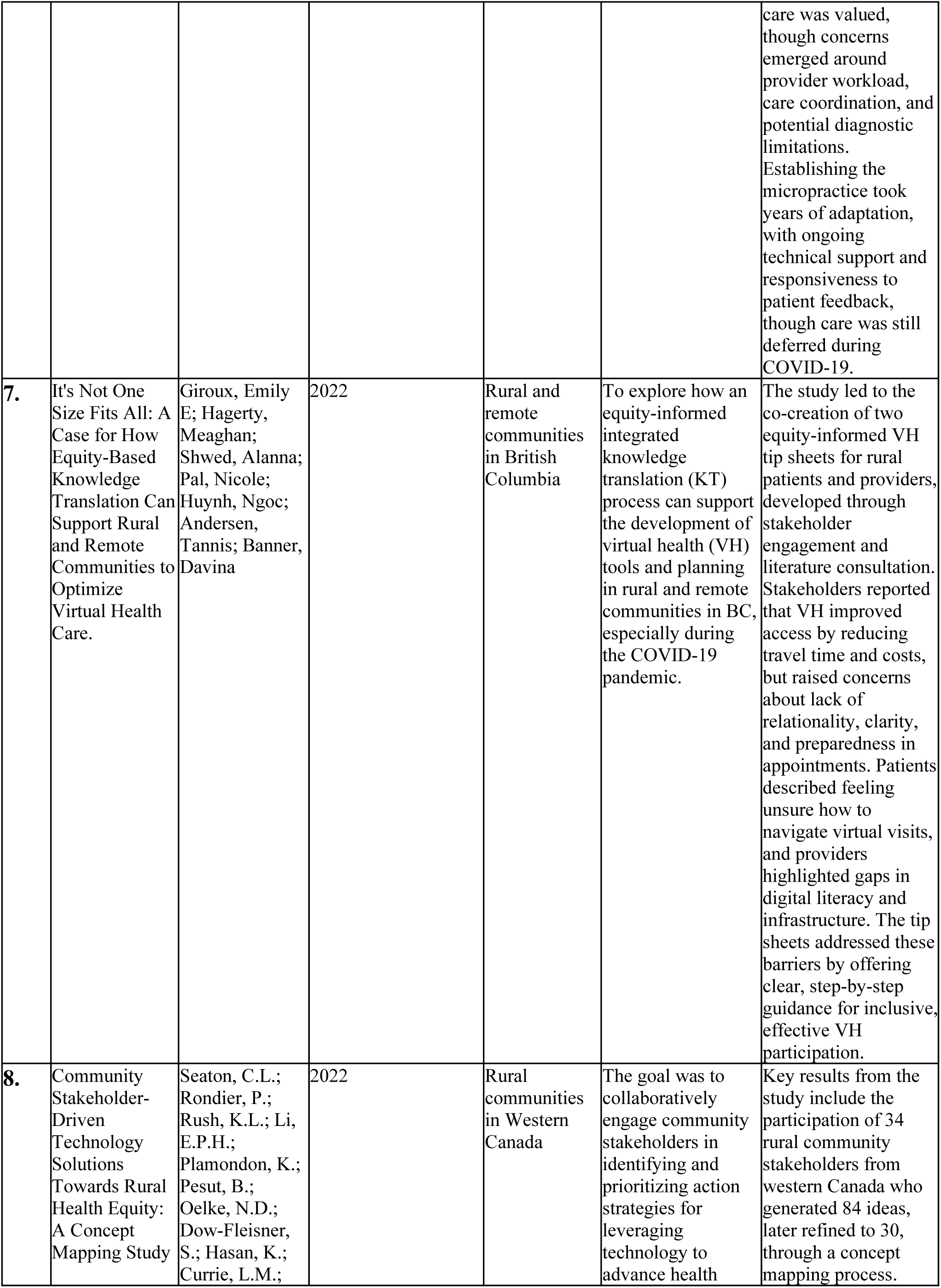
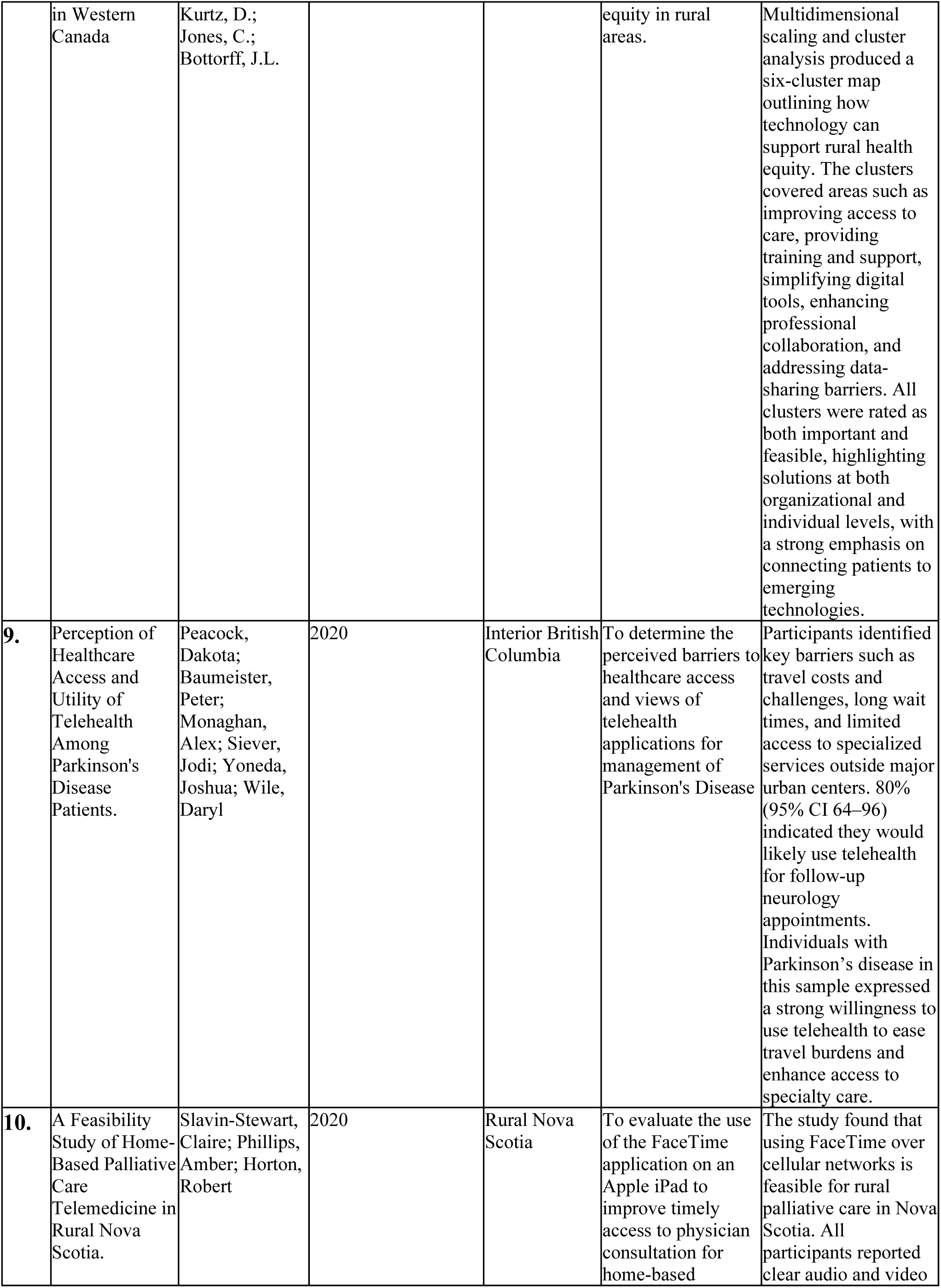
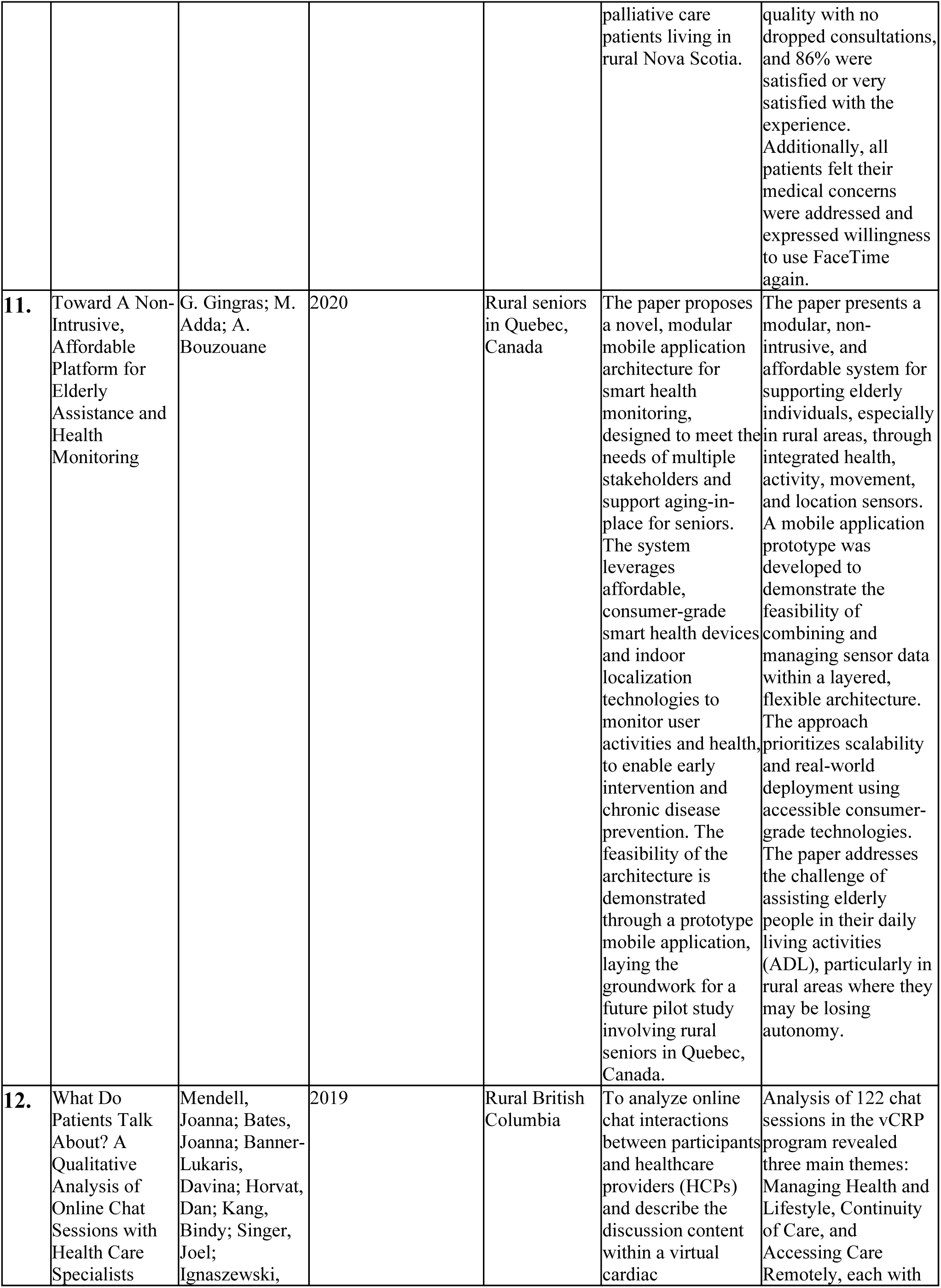
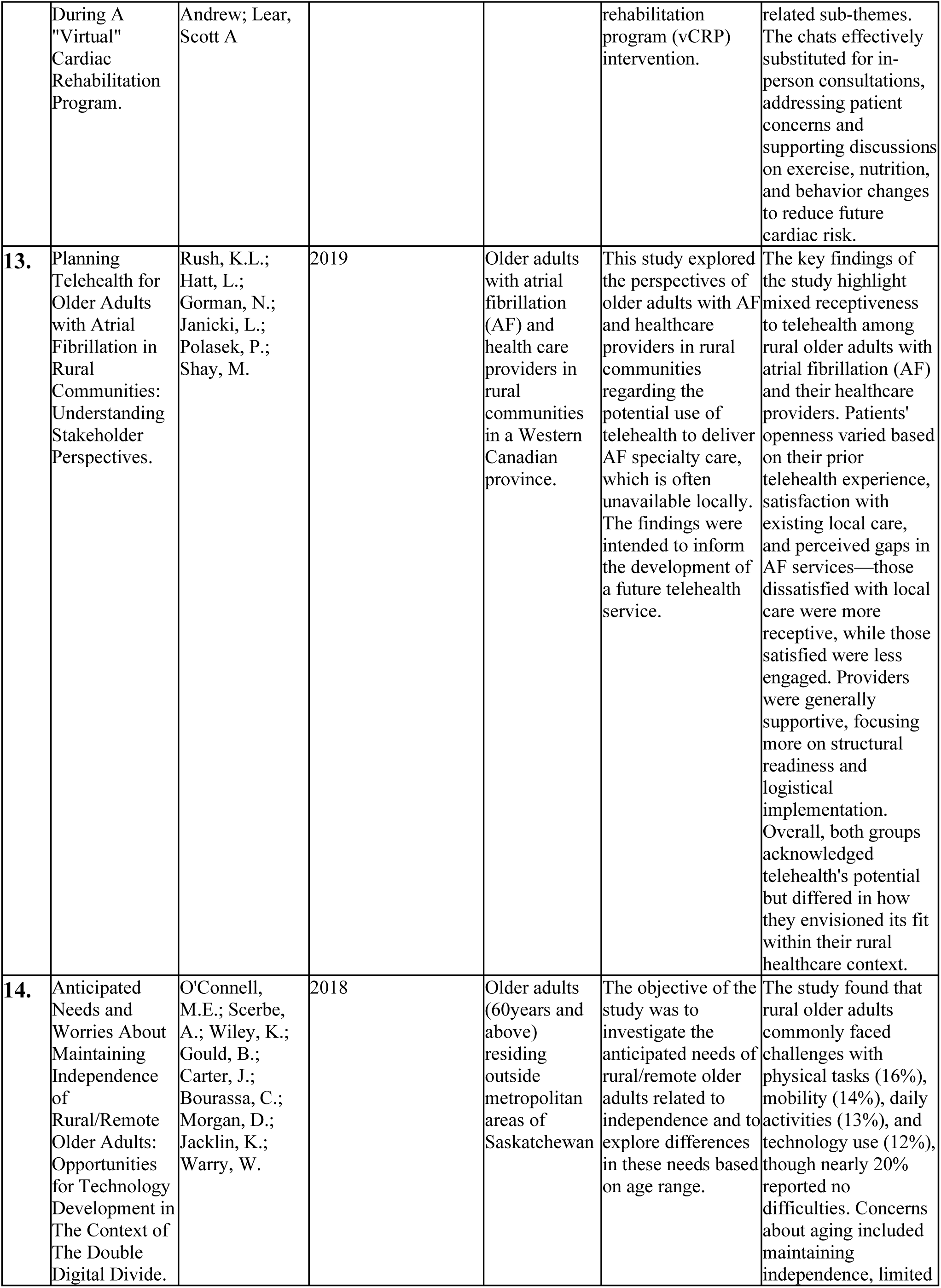
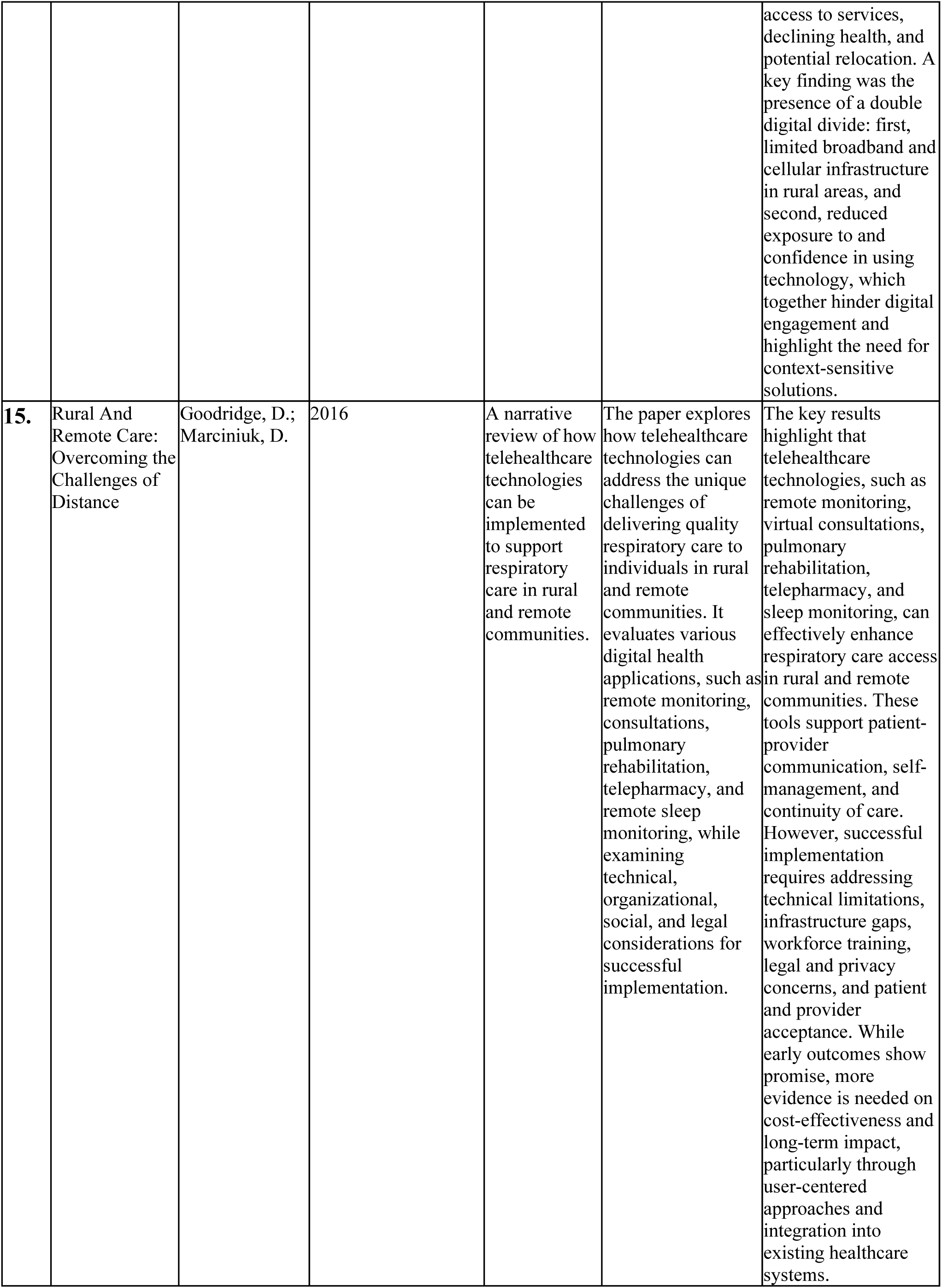
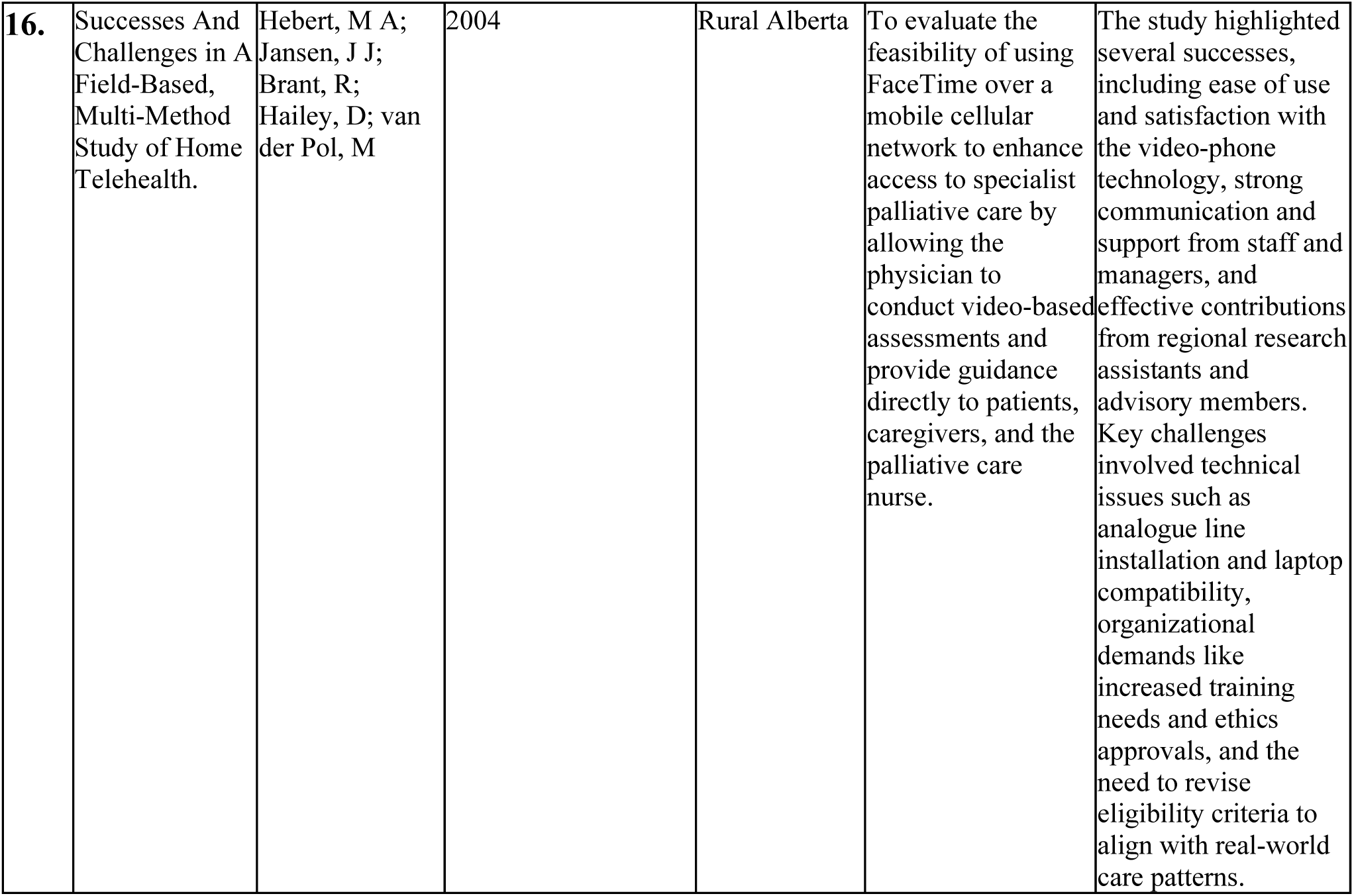
Summary of Data Extraction.

While this review excluded grey literature to maintain methodological transparency, we acknowledge that this may have limited the representation of relevant Indigenous-led or community-based digital health initiatives that may not appear in peer-reviewed databases.

## 3 Results

### 3.1 Study Characteristics

A total of sixteen studies met the inclusion criteria for this review. Table 1 presents the complete data extraction results, while Table 2 provides an overview of the search results. Most studies (n = 15) were conducted in rural regions of Canada. Among these articles, ten included a qualitative component, while only two were purely quantitative. Two articles, one a narrative review and the other a Prototype Development Paper, did not enroll patients but still provided relevant insights for our objectives. Most of the articles (n = 11) were published within the last five years, with all but one published between 2016 and 2024. One article from 2004 was included because of its direct relevance to our review questions.

**Table 2:** Overview of search results.

| Variable |  | Number |
| --- | --- | --- |
| Canadian province/study location | British Columbia | 5 |
|  | Ontario | 4 |
|  | Western Province | 2 |
|  | Nova Scotia | 1 |
|  | Alberta | 1 |
|  | Quebec | 1 |
|  | Saskatchewan | 1 |
|  | Review/study location not mentioned | 1 |
| Study Design | Mixed methods | 3 |
|  | Multi-method | 2 |
|  | Qualitative | 5 |
|  | Quantitative | 2 |
|  | Commentary | 1 |
|  | Narrative review | 1 |
|  | Prototype development Paper | 1 |
|  | Concept mapping | 1 |
| Year | 2024 | 3 |
|  | 2023 | 2 |
|  | 2022 | 3 |
|  | 2020 | 3 |
|  | 2019 | 2 |
|  | 2018 | 1 |
|  | 2016 | 1 |
|  | 2004 | 1 |

In what follows, we summarize the key findings, organized according to the four themes that align with the guiding research questions: (1) Readiness Tools, Frameworks, and Indicators; (2) Patient and Provider Perspectives; (3) Barriers and Corresponding Strategies; and (4) Outcomes and Impacts.

### 3.2 Readiness Tools, Frameworks, or Indicators

Despite a growing interest in digital health implementation in rural Canada, few studies employed formal tools or frameworks to assess community or patient readiness before rollout. Instead, researchers often inferred readiness through contextual factors or observed indirectly during implementation. Consistent indicators for readiness across several studies were the importance of digital infrastructure, user comfort with technology, and the adaptability of healthcare systems or providers. For instance, Burton et al. described how a micropractice took five years to optimize its virtual care delivery model, with success relying heavily on patient feedback, technical support, and provider autonomy (16). Other informal indicators of readiness included patients’ familiarity with digital tools, their willingness to engage actively in care, and provider comfort in adapting technological advancements to their clinical workflows.

Some studies underscored the *absence* of formal readiness assessment processes, highlighting a significant gap in implementation planning. For example, Fitzsimon et al. (2023) described the VTAC that was implemented within 12 days in response to COVID-19, without any structured evaluation of community preparedness (17). Similarly, Kornelsen et al. and Rouleau et al. reported virtual care implementations that proceeded without standardized tools (18,19). Both studies showed the importance of healthcare provider confidence with technology as well as the ease and comfort patients feel when communicating with healthcare providers through video or telephone. These factors are considered implicit readiness factors, especially when compassionate care delivery is involved (19).

Other papers proposed proxy readiness measures. For example, Giroux et al.used equity-informed integrated knowledge translation (KT) processes were used to identify community needs, infrastructural limitations, and key stakeholders (20). Rather than applying a traditional readiness framework, the study engaged rural and Indigenous community members to co-develop virtual health tools, using indicators such as digital literacy, broadband access, and community engagement to shape implementation. Petrie et al. took a similar community-driven approach and emphasized long-term relationship-building and cultural adaptation, such as hiring local coordinators and translating materials into Indigenous languages, as core readiness enablers (21).

Several studies highlighted the potential for using feasibility metrics as readiness proxies. For example, Salvin-Stewart et al. found that technical stability, patient satisfaction with audio/video quality, and willingness to reuse the service served as reliable indicators of virtual palliative care readiness (22). Peacock et al. also pointed to high patient comfort with videoconferencing and widespread access to telehealth infrastructure across 40 regional hospitals as signs of preparedness for neurology follow-up via telehealth (23).

Design-centered strategies develop technology solutions tailored to the realities of rural areas. For example, Gingras et al., took an implicit readiness-based approach, adapting solutions for low-tech rural settings by prioritizing simplicity, comfort, and affordability (24). Similarly, Mendell et al. showed that inclusion and exclusion criteria, such as requiring internet access and digital literacy, effectively filtered participants based on implicit readiness. However, the patients’ low uptake of chat sessions suggested that there were unmeasured barriers related to comfort with technology (25).

A few studies referenced existing models to analyze telehealth readiness. For example, Rush et al. applied the telehealth readiness framework developed by Jennett et al.’s (2003) to categorize users into different types, such as “genuinely ready” and “non-ready”. Their findings showed how patient satisfaction with rural healthcare influenced openness to virtual services (26). O’Connell et al. (2018) advocated for the Technology Acceptance Model (“rural TAM”) that integrates contextual rural factors like geographic isolation, infrastructure gaps, and the double digital divide (27). This framework, alongside the Matching Person and Technology Model, highlights the importance of aligning technology with users’ goals, environments, and perceived usefulness.

Finally, Goodridge and Marciniuk offered a multi-dimensional lens on readiness, identifying technical (infrastructure), organizational (staffing, integration), social (trust, digital literacy), and legal (privacy, data governance) factors as key domains (28). These dimensions, while not packaged as a formal tool, serve as a checklist for assessing digital health readiness in rural communities.

Overall, the existing literature highlights a significant lack of standardized tools for assessing readiness, despite a rich set of existing de facto indicators. These indicators include factors such as technological infrastructure, digital literacy, cultural compatibility, provider adaptability, and community engagement. Examples from these studies suggest a need to formalize and standardize readiness assessment practices in rural digital health implementations. Developing tailored, practical, equity-focused frameworks and tools could enhance planning and sustainability.

### 3.3 Patient and Provider Perspectives

Patients and providers in rural Canada have generally demonstrated positive attitudes toward home-based digital health technologies, although their reported attitudes are often highly context-dependent. In a study of a micropractice model, Burton et al. reported that patients felt empowered, less stressed, and more engaged in their care, while providers expressed pride in offering accessible, patient-centered services (16). Similarly, Fitzsimon et al. (2023) found that physicians appreciated the ability to reach underserved patients and described a sense of personal fulfillment, though some patients, particularly older adults, struggled with digital literacy (17). High patient satisfaction was also observed by Kornelsen et al., with 84 percent of patients recommending virtual surgical consults, though in-person visits remained preferred for emotionally significant care like surgery (18). In a culturally grounded remote monitoring initiative, Petrie et al. found universal patient satisfaction, with many reporting greater confidence in managing their health, and providers overwhelmingly supporting the program (21).

Provider perspectives were consistently supportive but tempered by concerns around system limitations. Providers also noted challenges in maintaining work-life boundaries when providing virtual care (16). Rouleau et al. described how nurses valued video visits to deliver compassionate care, though some felt digital formats constrained relational depth (19). Giroux et al. echoed this sentiment, with providers acknowledging the benefits of virtual care but expressing reservations about its adequacy as a full replacement for in-person services, especially when infrastructure and training were lacking (20). Hebert et al. found that nurses became more enthusiastic post-training, despite initial anxiety, indicating that onboarding and capacity-building are critical to long-term engagement (29). Meanwhile, Goodridge and Marciniuk documented provider worries about increased workload, disrupted workflows, and legal complexities, highlighting the importance of organizational support and clear role definitions (28). Some examples of the legal concerns mentioned include challenges related to patient privacy, securing informed consent, and maintaining confidentiality in the transfer, storage, and sharing of medical data, alongside issues surrounding provider authentication and liability.

On the patient side, virtual care was often associated with convenience, empowerment, and reduced logistical burdens. For example, Peacock et al. noted that patients appreciated telehealth’s efficiency, especially for follow-ups, and valued the reduced stress of avoiding travel (2). In a virtual cardiac rehabilitation program, Mendell et al. reported that participants enjoyed the motivational aspects of digital interaction, though one user preferred phone calls over chat due to usability frustrations (25). Kornelsen et al. provided virtual consultations to patients via videoconference, with phone consultations acting as a backup option to resolve connectivity issues (18). These examples suggest that flexible communication formats can enhance engagement.

Seaton et al. found that patients favored user-friendly tools supported by human assistance, emphasizing that simplicity and reliability often outweighed the benefits of innovation (30). In contrast, Rush et al. observed mixed attitudes: patients who were dissatisfied with existing services showed greater openness to virtual care, while those content with current access were more resistant to change (26). O’Connell et al. highlighted a similar nuanced range of attitudes among older adults, who expressed both anxiety and resilience when navigating technology (27). This ambivalence reflected the tension between discomfort with unfamiliar digital tools and a demonstrated willingness to adapt. Their responses were shaped by perceived usefulness of the technology, confidence in their own ability to use it (self-efficacy), and access to support systems, such as assistance from family or community-based resources, that made engagement with technology more feasible.

Despite overall enthusiasm for home-based digital care, several concerns affected patient attitudes. Giroux et al. noted that patients missed relational depth and emotional clarity in virtual visits, underscoring the importance of relational care elements in digital settings (20). Goodridge and Marciniuk found that some patients feared technical failure, over-reliance on clinicians, and the loss of in-person interactions (28). Likewise, Seaton et al. identified connectivity and complexity as barriers, reinforcing the need for strong infrastructure and intuitive design (30).

Notably, a few studies, including those by Fitzsimon et al. (2023) and Gingras et al., did not yet collect direct feedback on user attitudes, though future data collection was planned (6,17). This highlights a broader critical need across the field for consistent integration of user experience metrics in digital health evaluations. Ongoing measurement of patient and provider experiences is essential for continuous improvement.

In summary, attitudes toward digital health in rural Canada are most favorable when technologies are accessible, supportive, and designed with empathy and practical needs in mind. Successful implementation depends on co-design with patients and providers, developing patient-centered digital solutions, ongoing training, and the inclusion of emotional and relational components. Flexibility in delivery formats and continuous evaluation of user satisfaction can further strengthen adoption and sustained use.

### 3.4 Barriers and Corresponding Strategies

The implementation of home-based digital health technologies in rural Canada faces a complex web of interrelated barriers across various levels, including technological, organizational, social, and regulatory domains.

#### 3.4.1 Technological domain

Connectivity issues resulting from technological and infrastructural challenges were common. Several studies highlighted that low-quality broadband and cellular access frequently disrupted virtual visits, while unstable power supplies further complicated digital service delivery in remote areas (17,18,28). Seaton et al. also noted that even when digital tools are available, gaps in connectivity limit their effectiveness (30). To address these challenges, several strategies have been proposed. These include using low-bandwidth access points such as toll-free telephone lines (17), deploying reliable cellular-based platforms (22), and implementing localized data processing to reduce dependence on cloud-based systems and improve speed (6).

Hardware and device limitations also pose significant barriers, especially for older adults. Gingras et al. and Petrie et al. reported that high costs, complex authentication protocols, and limited compatibility with existing devices can reduce access (6,21). Some strategies suggested include the use of simplified authentication process and password reset functions to accommodate seniors.

Digital literacy remains a core challenge across patient and provider populations. O’Connell et al., Peacock et al., and Rush et al. found that many older adults lacked the confidence or familiarity needed to engage with telehealth platforms (2,21,26). For providers, unfamiliarity with digital systems could translate to increased stress, as noted by Rouleau et al., Hebert et al., and Burton et al., who described workflow strain, blurred professional boundaries, and burnout (16,19,29). Effective strategies included remote training programs, intuitive system design, repetitive demonstrations, and workflow reconfiguration to ensure digital care responsibilities are sustainable.

#### 3.4.2 Organizational domain

At the system level, interoperability issues and workforce constraints hinder scale-up. For example, a lack of electronic medical record (EMR) integration between organizations limits care continuity (16,21,30). To mitigate this, teams employed communication protocols, bridging platforms like the “Connecting Ontario” portal (Petrie et al.), and patient-facing tools to coordinate follow-up (21). Human resource limitations, including underestimated training needs and a shortage of tech-competent staff, were prominent in some studies (29,30). These were countered by creating “tech ambassador” roles, targeted onboarding, and flexible recruitment strategies tailored to rural staffing realities.

#### 3.4.3 Social Domain

Cultural, relational, and emotional barriers also influence digital health adoption. Some studies found that digital formats can reduce the emotional nuance and relational depth of care (19,20). For example, patients missed the non-verbal cues and physical presence of in-person visits, particularly in emotionally significant contexts like surgery. To address this, providers emphasized the use of video over audio-only formats, incorporated hybrid care models, and developed training to support “relational care” in digital settings. Petrie et al. and Goodridge & Marciniuk also highlighted cultural mismatches and language barriers, especially in Indigenous communities (21,28). Culturally tailored materials, translated communications, and community leaders’ involvement were effective in overcoming these challenges.

Adoption can also hindered by patient hesitancy, low engagement, and digital skepticism. For example, Mendell et al. and Kornelsen et al. described how chat-based platforms experienced low uptake, particularly among users with basic digital skills (18,25). Some patients expressed a preference for phone or in-person communication. Strategies to improve engagement include offering multiple communication formats (video, chat, phone), guided onboarding, and fallback options for those unable or unwilling to use digital platforms. Studies by Rush et al. and O’Connell et al. revealed skepticism among patients who were satisfied with their current care or unsure of digital health’s value (26,27). In these cases, trust-building efforts, co-design initiatives, and patient-centered education were key to shifting attitudes.

#### 3.4.4 Regulatory Domain

Legal, ethical, and regulatory concerns further complicate virtual care delivery. For example, Goodridge & Marciniuk and Rouleau et al. noted that privacy risks and liability fears affected provider confidence, particularly in remote assessments (19,28).

Regulatory issues, such as difficulties prescribing controlled substances or ordering labs remotely, were noted by Fitzsimon et al (17). Some solutions they identified included secure platforms, clearly defined virtual care policies, and collaborations with local clinical teams or paramedics to bridge physical service gaps.

#### 3.4.5 Fundamental strategies to overcome barriers

In response to these multi-level barriers, several overarching strategies emerged. Giroux et al. demonstrated the value of equity-informed implementation using integrated knowledge translation (KT) approaches, including co-designed materials in plain language (20). Seaton et al. and Peacock et al. emphasized the role of trusted health workers, such as nurses and paramedics, in bridging tech gaps and facilitating digital assessments. Fitzsimon et al. and Slavin-Stewart et al. advocated for low-tech access points like toll-free numbers and consumer tools (e.g., FaceTime on iPads) to improve usability (17,22). Finally, Goodridge & Marciniuk and O’Connell et al. called for systemic investment in infrastructure, workforce development, and policy frameworks to ensure that digital health initiatives are not only implemented but sustained and scaled appropriately for rural populations (27,28).

### 3.5 Outcomes and Impacts of Home-Based Digital Health Technologies

This section summarizes the reported outcomes and impacts of home-based digital health technologies in rural Canada, as documented across the included studies. Findings are presented at the patient, provider, and health system levels, as well as feasibility.

#### 3.5.1 Patient-Level Outcomes

Across multiple studies, patients reported positive experiences with home-based digital health technologies, particularly concerning empowerment, self-management, and access. For example, in a virtual micropractice model, Burton et al. observed that patients felt more in control of their health and were less anxious about care continuity during the COVID-19 pandemic (16). Similarly, in a culturally grounded heart failure monitoring program, Petrie et al. found that 93 percent of eligible patients were successfully titrated to optimized guideline-directed therapy, and 87 percent would recommend the program (21). These outcomes were tied to strong adherence rates and perceived improvements in disease self-management. In a virtual cardiac rehabilitation program, Mendell et al. highlighted gains in patient knowledge, motivation, and behavioral change, including improved diets and exercise routines (25).

Patient satisfaction with digital care was also high. Slavin-Stewart et al. reported 100 percent satisfaction with home-based palliative consultations, and Kornelsen et al. found that virtual surgical consultations scored an average of 8.38/10 (18,22). Patients cited substantial time and cost savings, with 36 percent saving over CAD250 and many avoiding long travel times. Access to virtual consultations sometimes prevented patients from forgoing or delaying essential procedures. Anticipated access gains were especially noted by Peacock et al. and Rush et al., who emphasized benefits for mobility-constrained individuals and those living far from specialist services (2,26). While some studies, such as Giroux et al. and Seaton et al., did not measure outcomes directly, they discussed the expected equity impacts of culturally tailored, co-developed digital tools (20,30).

#### 3.5.2 Provider-Level Outcomes

Providers experienced a range of outcomes, both positive and challenging. Fitzsimon et al. noted that rural physicians experienced increased professional fulfillment from reaching underserved populations and improved their remote assessment and history-taking skills (17). Similarly, Rouleau et al. found that nurses gained valuable insights into patients’ home environments, which improved the delivery of contextual care (19). However, researchers found that high workloads, stress, and burnout were key risks to sustaining compassionate virtual care.

In some instances, communication and care coordination also improved. Burton et al. reported more timely exchanges using asynchronous messaging and EMR portals (16). Petrie et al. described how coordinated care was improved through EMR workarounds and patient coaching by local staff, demonstrating the importance of flexible workflows and cross-system collaboration (21).

#### 3.5.3 Health System and Population-Level Outcomes

Several studies documented system-level improvements, particularly around the use and efficiency of emergency care. The VTAC hybrid care model reported a 32.9% reduction in low-acuity emergency department visits (17). Simultaneously, there was an increase in high-acuity visits and hospitalizations, suggesting better triage and earlier intervention. Additionally, health system spending in VTAC regions grew at a slower rate compared to neighboring jurisdictions. Another study found that remote programs for chronic respiratory disease with home-based pulmonary rehabilitation effectively reduced emergency department use and matched the effectiveness of in-clinic programs (28).

Economic gains were observed not only in overall system-wide savings but also for patients. Kornelsen et al. emphasized substantial time and financial savings for patients. Goodridge & Marciniuk described how remote care, particularly involving robotic telepresence, helped reduce costly medical evacuations by up to 60 percent in northern Indigenous communities (18,28). These developments suggest the potential for early cost-effectiveness and sustainability.

#### 3.5.4 Feasibility and Scalability

Evidence from several pilot studies supports the technical and operational feasibility of scaling home-based digital care in rural contexts. For example, Slavin-Stewart et al. demonstrated that mobile video consultations over LTE networks could be delivered consistently and with high patient satisfaction in rural Nova Scotia. Petrie et al. exceeded enrollment targets in their heart failure monitoring program, which they attributed to strong infrastructure, community partnerships, and cultural tailoring (21). Although outcome data from older studies, such as those from Hebert et al., are still emerging, qualitative findings support the feasibility of these approaches (29). Gingras et al. similarly discussed projected benefits, including earlier detection of anomalies and prolonged independent living for patients; however, these remain theoretical until their pilot testing is completed (24). Overall, the findings suggest promising potential for low-tech, flexible home-based digital health technologies to enhance care delivery in rural settings while also highlighting the need for continuous evaluation to determine long-term viability and adaptation to meet community-specific needs.

## 4 Discussion

This scoping review highlights a growing body of evidence supporting the use of home-based digital health technologies in rural Canada. It focuses on readiness (both community and practitioner), stakeholders’ perspectives, barriers to implementation, strategies to overcome these challenges, and the observed outcomes and impacts. Overall, findings demonstrate strong patient satisfaction, perceived healthcare access and quality improvements, and early indications of system-level benefits. However, existing literature also shows important challenges and limitations, particularly related to the limited use of formal readiness assessment tools, poor infrastructure, digital literacy, and long-term sustainability. Addressing these issues is essential to ensure equitable and effective implementation.

### 4.1 Readiness Tools and Frameworks

Few used formal readiness tools before implementing digital health technologies in rural Canada. Instead, researchers often assessed readiness was often assessed through contextual observations, such as broadband access, digital literacy, and community engagement. For example, successful programs frequently relied on informal indicators like user comfort with technology, provider adaptability, or feasibility metrics, including video and audio quality satisfaction, and people’s to re-use the technology. Only a small number applied structured models like Jennett et al.’s telehealth readiness framework or the Technology Acceptance Model (26,27). Some models emphasized co-developed, equity-informed planning approaches.

Community-led programs, such as those by Petrie et al., Giroux et al. incorporated readiness factors like infrastructure fit, cultural adaptation, and stakeholder relationships, even though these factors were not explicitly labeled as readiness (20,21). These findings indicate a critical gap in standardized, equity-oriented readiness tools tailored to rural and Indigenous contexts.

In Canada, Indigenous-led initiatives, including British Columbia’s Rural, Remote, First Nations and Indigenous COVID-19 Response Framework (31) demonstrate promising models rooted in cultural safety and local governance. Internationally, frameworks such as Australia’s National Digital Health Strategy (32) and the U.S. National Quality Forum’s Rural Telehealth Readiness Framework (33) similarly prioritizes infrastructure, digital inclusion, and responsive design. These examples reinforce the value of tailoring readiness tools to the sociocultural and structural realities of underserved populations. Developing and formalizing such tools could strengthen planning, alignment, and evaluation processes. More importantly, ensuring that rural ehealth interventions use these frameworks before implementation will improve their expected outcomes.

### 4.2 Patient and Provider Perspectives

Overall, patients and providers generally expressed positive attitudes toward home-based digital health in rural Canada, although their responses were context dependent. Patients frequently reported increased empowerment, improved chronic disease management, and reduced logistical burdens. Culturally grounded models further enhanced trust, health literacy, and confidence. However, uncertainty persisted, especially among older adults, due to perceived usefulness, patient self-efficacy, and the availability of support from family or communities. These outcomes are similar to findings from rural digital health programs in Australia and the U.S., where virtual care improved autonomy and access (34–36).

Providers’ outcomes were more mixed. While some studies highlighted improved communication, diagnostic skills, professional fulfillment, and workflow efficiency (Fitzsimon et al., 2023; Burton et al., 2022), others raised concerns about burnout, blurred role boundaries, and the emotional strain of maintaining compassionate care in virtual formats, especially where digital infrastructure, staffing, or training was inadequate (Rouleau et al., 2024). Hybrid delivery options helped support both relational and technological aspects of healthcare challenges. Satisfaction was highest when technologies were flexible, user-friendly, and supported by human assistance. Offering communication options, video, phone, or chat, and in-person visits helped accommodate individual preferences and mitigated usability or connectivity challenges. These patterns were also documented in US, Australian, and New Zealand studies (37–39).

Enhanced care continuity was reported in studies where digital tools were integrated with EMRs and supported by patient coaching or local staff (Burton et al., 2022; Petrie et al., 2024). One study suggested that hybrid models that begin with an initial in-person visit followed by virtual follow-ups improved therapeutic relationships among indigenous communities (40). These models suggest that digital health can complement, rather than replace, existing care structures. Without flexibility, system-level support, and clearly defined roles, the sustainability of digital health among rural providers may be compromised despite providers’ initial enthusiasm.

Remarkably, some studies had not yet collected direct user feedback, reflecting a broader limitation in the depth of insight into user experiences. Consistent integration of patient and provider experience metrics remains a critical gap and priority for future research.

The insights emphasize that sustainable adoption depends not only on co-design with users before and during implementation but also on investments in digital literacy, emotional support, and workload balance. Policymakers and health systems should prioritize flexible delivery models, inclusive evaluation practices, and provider well-being to promote equitable and lasting digital health integration in rural settings.

### 4.3 Barriers and Corresponding Strategies

Technological barriers are among the most frequently reported challenges in delivering home-base digital health. In many remote areas, poor broadband and cellular coverage often disrupt care delivery. Additionally, hardware and device limitations, such as high costs, complex authentication protocols, and incompatibility with existing technology, disproportionately affect older adults and low-income users (6,21). These findings align with experiences in rural Australia, the United States, and other high-income countries where gaps in digital infrastructure and affordability constraints similarly hinder virtual care uptake (Bradford et al., 2016; Harkey et al., 2020; Maita et al., 2024b). In response, effective mitigation strategies have included the use of low-bandwidth tools like toll-free telephone lines, simplified consumer-grade equipment, and local data processing solutions (e.g., edge computing). Some studies advocated using simplified, consumer-grade equipment and low-cost solutions, often combined with patient coaching or staff support to help with setup and daily use. Digital exclusion risks were mitigated through training and equipment loans (41,42).

Digital literacy and user confidence are critical issues impacting both patients and providers. Many older adults lack familiarity with telehealth platforms, while numerous providers report stress and burnout related to unfamiliar digital workflows and blurred professional boundaries (19,27,29). Training programs, intuitive system designs, and role-specific support, such as “tech ambassadors”, have been identified as effective means to enhance uptake and reduce patients’ cognitive load.

At the organizational level, challenges include poor electronic medical record (EMR) integration between institutions, a lack of cross-platform communication, and workforce shortages. Interventions like shared digital portals (e.g., Connecting Ontario), patient-facing tracking tools, and targeted onboarding for rural clinicians have helped to alleviate these constraints (16,21) However, workforce gaps remain a persistent issue across high-income rural health systems (37).

Social and cultural barriers also play a significant role, particularly in Indigenous and underserved communities. Patients often report a diminished emotional connection in digital formats, especially in sensitive contexts like surgical care. Some programs have implemented hybrid models that combine in-person visits with follow-up telehealth sessions to strengthen trust and relational continuity (40). Addressing cultural mismatches, language barriers, and institutional distrust has involved strategies such as co-design, the use of local coordinators, and the translation of materials into Indigenous languages (Petrie et al., 2024; Giroux et al., 2022). Similar strategies have been effective in Indigenous communities in Australia, where co-developed and culturally tailored telehealth models have improved engagement (36). However, one study showed that 40 percent of the 321 studies included in the review did not report any Indigenous involvement in telehealth services or research processes. This highlights the need for standards ensuring consistent patient involvement in underserved populations.

Regulatory and legal issues, such as concerns surrounding privacy, liability, and limitations in prescribing or diagnostic authority, further complicate implementation efforts. Secure platforms, virtual care policies, and collaborations with paramedics or local clinics have helped navigate these constraints (Goodridge & Marciniuk, 2016; Rouleau et al., 2024).

Despite promising implementation strategies, structured evaluations of implementation are often lacking. Very few studies have applied formal frameworks to assess acceptability, feasibility, or fidelity, which limits insights into what works and under what conditions. To strengthen future implementations, it is recommended to use theory-driven frameworks such as the Consolidated Framework for Implementation Research (CFIR) or RE-AIM. These tools can support the design and evaluation of interventions by addressing factors such as stakeholder engagement, contextual adaptation, and sustainability.

In summary, while various practical strategies are being employed to tackle the complex barriers to rural digital health implementation, future efforts should adopt a more systematic and equity-informed approach. Integrating implementation science and paying closer attention to both patients’ and providers’ lived realities, especially those in marginalized settings, will be essential for achieving long-term success.

### 4.4 Outcomes and Impacts of Home-Based Digital Health Technologies

The reviewed studies consistently reported positive outcomes across patient, provider, and system levels. When it comes to using digital health technologies in their homes, patients in rural Canada experienced high satisfaction, greater self-management, and reduced travel-related burdens, particularly in chronic disease (e.g., cardiac rehab, palliative care) and culturally grounded programs. These results confirm findings from rural programs in Australia and the U.S., where virtual care enhanced patients’ autonomy, reduced missed appointments, and supported care continuity in underserved communities (34–36).

Providers reported improved communication and skill development, but many also faced burnout, role strain, and emotional fatigue, especially when digital infrastructure, staffing, or training were inadequate. These tensions have also been documented internationally, reinforcing the need for flexible care delivery models, team-based support, and clear role boundaries (38).

At the system level, well-integrated digital health interventions helped reduce low-acuity emergency department visits and slow health system cost growth. Programs like VTAC demonstrated more effective triage and early intervention. Similar patterns were noted in respiratory disease programs, where remote care led to fewer hospitalizations without compromising clinical outcomes (28). Such patterns align with digital health models in other high-income countries that have shifted care from reactive to preventive modes while containing costs. This indicates that deploying digital tools within supportive system frameworks can help reduce the burden of care in acute settings.

Despite these benefits, outcome measurement across the literature remains inconsistent. Many studies were exploratory and lacked follow-up. To strengthen evidence for scale-up, future research should incorporate structured evaluation frameworks like CFIR or RE-AIM, focusing on sustainability, equity, and patient-centered metrics such as quality of life.

### 4.5 Implementation Considerations and Future Directions

To maximize the impact of digital health in rural Canada, implementation strategies must be tailored to the realities of these underserved communities. This includes ensuring reliable broadband or cellular infrastructure, addressing digital literacy for both patients and providers, and designing tools that are intuitive and culturally appropriate (6,21,30). Multiple studies emphasized the value of co-design, community engagement, and human-centered support systems (e.g., tech ambassadors, nurse coordinators, paramedics) as essential to uptake and sustainability (16,20).

Efforts to implement digital health initiatives must take into account the broader social determinants of health. This includes factors such as trust in institutions, language barriers, and historical inequities, especially in Indigenous and remote communities. To prevent these initiatives from exacerbating existing disparities, it is essential to focus on equity-based planning and integrated knowledge translation, which can help advance health justice (20,27).

Furthermore, it is crucial to prioritize the regular collection of user feedback and to apply implementation science methods to assess scalability, cost-effectiveness, and contextual success factors. Future research should aim to generate high-quality, long-term evidence, particularly regarding clinical outcomes, economic impacts, and sustainability in diverse rural and remote settings.

### 4.6 Limitations of This Review

This review was limited to peer-reviewed, English-language publications. As such, it may have excluded relevant grey literature, policy documents, or Indigenous-led initiatives not captured in academic databases. Publication bias may also have led to overrepresentation of well-resourced or successful interventions.

Despite promising results, many studies remain limited in their ability to assess long-term impacts. Several, including Giroux et al., Seaton et al., Rush et al., and O’Connell et al., focused on anticipated rather than measured outcomes (20,27,30). These conceptual or planning-phase studies highlighted the potential of digital tools but emphasized the need for robust, longitudinal evaluations. Hebert et al. also reported difficulties in collecting real-world economic data, suggesting a broader need for implementation science methods that capture cost-effectiveness and quality-of-life improvements in rural contexts (29).

## 5 Conclusion

This review summarizes emerging evidence on the implementation and outcomes of home-based digital health technologies in rural Canada. Clear benefits include increased patient empowerment, improved access to care, and enhanced system efficiency. Providers expressed enthusiasm, improved skills, and noted some structural strain regarding the use of these technologies. To sustain and scale these benefits, implementation must be grounded in the specific context of rural communities, informed by equity, and supported by adequate infrastructure, policy, and ongoing evaluation. Digital solutions should go beyond simply providing access; they must focus on achieving long-term sustainability, promoting relational care, and ensuring structural inclusion that addresses the unique needs of rural populations. Future efforts should prioritize rigorous evaluations, align with policy frameworks, and incorporate marginalized voices in the design and governance of digital health systems.

## Data Availability

All data produced in the present work are contained in the manuscript.

## 7 Author Contributions

JL: Writing – original draft, Writing – review & editing, Conceptualization, Methodology, Formal analysis, Project administration; AS: Writing – original draft, Writing – review & editing, Conceptualization, Methodology, Formal analysis, Project administration; GO: Writing – original draft, Writing – review & editing, Conceptualization, Methodology, Formal analysis, Project administration.

## 9 Acknowledgments

The authors gratefully acknowledge Gotcare and Quinte Health for their collaboration on the broader project within which this scoping review was undertaken. We also extend our sincere thanks to Eden Kinzel, Liaison & Education Librarian, for her guidance in developing a rigorous review process. We are deeply grateful to Enid Montague and Gonzalo Romero, who served as faculty mentors and provided invaluable guidance and support throughout this research. This work was supported through the Reach Alliance, a partnership between the University of Toronto’s Munk School of Global Affairs & Public Policy, and the Mastercard Centre for Inclusive Growth. This research is supported by the DIGITAL Innovation Cluster, with funding from Innovation, Science and Economic Development Canada.

## Reference styles

1. Wilson R, Rourke J, Oandasan IF, Bosco C. Progress made on access to rural health care in Canada. Can Fam Physician. 2020 Jan;66(1):31–6.

2. Peacock D, Baumeister P, Monaghan A, Siever J, Yoneda J, Wile D. Perception of Healthcare Access and Utility of Telehealth Among Parkinson’s Disease Patients. Canadian Journal of Neurological Sciences / Journal Canadien des Sciences Neurologiques. 2020 Sep 26;47(5):700– 4.

3. Syed ST, Gerber BS, Sharp LK. Traveling Towards Disease: Transportation Barriers to Health Care Access. J Community Health. 2013 Oct 31;38(5):976–93.

4. Cochran AL, McDonald NC, Prunkl L, Vinella-Brusher E, Wang J, Oluyede L, et al. Transportation barriers to care among frequent health care users during the COVID pandemic. BMC Public Health. 2022 Sep 20;22(1):1783.

5. Buyting R, Melville S, Chatur H, White CW, Légaré JF, Lutchmedial S, et al. Virtual Care With Digital Technologies for Rural Canadians Living With Cardiovascular Disease. CJC Open. 2022 Feb;4(2):133–47.

6. Gingras G, Adda M, Bouzouane A. Toward a Non-Intrusive, Affordable Platform for Elderly Assistance and Health Monitoring. In: 2020 IEEE 44th Annual Computers, Software, and Applications Conference (COMPSAC). IEEE; 2020. p. 699–704.

7. Maita KC, Maniaci MJ, Haider CR, Avila FR, Torres-Guzman RA, Borna S, et al. The Impact of Digital Health Solutions on Bridging the Health Care Gap in Rural Areas: A Scoping Review. Perm J. 2024 Sep 16;28(3):130–43.

8. Bouabida K, Chaves BG, Anane E, Jagram N. Navigating the landscape of remote patient monitoring in Canada: trends, challenges, and future directions. Front Digit Health. 2025 Feb 4;7.

9. O’Connell ME, Scerbe A, Wiley K, Gould B, Carter J, Bourassa C, et al. Anticipated needs and worries about maintaining independence of rural/remote older adults: Opportunities for technology development in the context of the double digital divide. Gerontechnology. 2018 Oct 8;17(3):126–38.

10. Wosny M, Strasser LM, Hastings J. Experience of Health Care Professionals Using Digital Tools in the Hospital: Qualitative Systematic Review. JMIR Hum Factors. 2023 Oct 17;10:e50357.

11. Tawfik DS, Sinha A, Bayati M, Adair KC, Shanafelt TD, Sexton JB, et al. Frustration With Technology and its Relation to Emotional Exhaustion Among Health Care Workers: Cross-sectional Observational Study. J Med Internet Res. 2021 Jul 6;23(7):e26817.

12. Arksey H, O’Malley L. Scoping studies: towards a methodological framework. Int J Soc Res Methodol. 2005 Feb;8(1):19–32.

13. Levac D, Colquhoun H, O’Brien KK. Scoping studies: advancing the methodology. Implementation Science. 2010 Dec 20;5(1):69.

14. Moher D, Liberati A, Tetzlaff J, Altman DG. Preferred Reporting Items for Systematic Reviews and Meta-Analyses: The PRISMA Statement. PLoS Med. 2009 Jul 21;6(7):e1000097.

15. Buyting R, Melville S, Chatur H, White CW, Légaré JF, Lutchmedial S, et al. Virtual Care With Digital Technologies for Rural Canadians Living With Cardiovascular Disease. CJC Open. 2022 Feb;4(2):133–47.

16. Burton L, Rush KL, Smith MA, Davis S, Rodriguez Echeverria P, Suazo Hidalgo L, et al. Empowering Patients Through Virtual Care Delivery: Qualitative Study With Micropractice Clinic Patients and Health Care Providers. JMIR Form Res. 2022 Apr 27;6(4):e32528.

17. Fitzsimon J, Patel K, Peixoto C, Belanger C. Family physicians’ experiences with an innovative, community-based, hybrid model of in-person and virtual care: a mixed-methods study. BMC Health Serv Res. 2023 Jun 3;23(1):573.

18. Kornelsen J, Taylor M, Ebert S, Skinner T, Stoll K. Rural patients’ experiences with anesthesia and surgical consultations in British Columbia: A survey-based comparison between virtual and in-person modalities. Digit Health. 2024 Jan 28;10.

19. Rouleau G, Wu K, Parry M, Richard L, Desveaux L. Providing compassionate care in a virtual context: Qualitative exploration of Canadian primary care nurses’ experiences. Digit Health. 2024 Jan 9;10.

20. Giroux, Hagerty, Shwed, Pal, Huynh, Andersen, et al. Its not one size fits all: a case for how equity-based knowledge translation can support rural and remote communities to optimize virtual health care. Rural Remote Health. 2022 May 9;

21. Petrie S, Ross H, Simard A, Innes E, Kioke S, Kozuszko S, et al. BRINGING CARE CLOSE TO HOME: REMOTE MANAGEMENT OF HEART FAILURE IN PARTNERSHIP WITH INDIGENOUS COMMUNITIES IN NORTHERN ONTARIO. Canadian Journal of Cardiology. 2023 Oct;39(10):S59.

22. Slavin-Stewart C, Phillips A, Horton R. A Feasibility Study of Home-Based Palliative Care Telemedicine in Rural Nova Scotia. J Palliat Med. 2020 Apr 1;23(4):548–51.

23. Peacock D, Baumeister P, Monaghan A, Siever J, Yoneda J, Wile D. Perception of Healthcare Access and Utility of Telehealth Among Parkinson’s Disease Patients. Canadian Journal of Neurological Sciences / Journal Canadien des Sciences Neurologiques. 2020 Sep 26;47(5):700– 4.

24. Gingras G, Adda M, Bouzouane A. Toward a Non-Intrusive, Affordable Platform for Elderly Assistance and Health Monitoring. In: 2020 IEEE 44th Annual Computers, Software, and Applications Conference (COMPSAC). IEEE; 2020. p. 699–704.

25. Mendell J, Bates J, Banner-Lukaris D, Horvat D, Kang B, Singer J, et al. What Do Patients Talk About? A Qualitative Analysis of Online Chat Sessions with Health Care Specialists During a “Virtual” Cardiac Rehabilitation Program. Telemedicine and e-Health. 2019 Jan;25(1):71–8.

26. Rush KL, Hatt L, Gorman N, Janicki L, Polasek P, Shay M. Planning Telehealth for Older Adults With Atrial Fibrillation in Rural Communities: Understanding Stakeholder Perspectives. Clin Nurs Res. 2019 Feb 20;28(2):130–49.

27. O’Connell ME, Scerbe A, Wiley K, Gould B, Carter J, Bourassa C, et al. Anticipated needs and worries about maintaining independence of rural/remote older adults: Opportunities for technology development in the context of the double digital divide. Gerontechnology. 2018 Oct 8;17(3):126–38.

28. Goodridge D, Marciniuk D. Rural and remote care. Chron Respir Dis. 2016 May 21;13(2):192– 203.

29. Hebert MA, Jansen JJ, Brant R, Hailey D, Van Der Pol M. Successes and challenges in a field-based, multi-method study of home telehealth. J Telemed Telecare. 2004 Nov 1;10(1_suppl):41–4.

30. Seaton CL, Rondier P, Rush KL, Li EPH, Plamondon K, Pesut B, et al. Community stakeholder-driven technology solutions towards rural health equity: A concept mapping study in Western Canada. Health Expectations. 2022 Dec 17;25(6):3202–14.

31. Government of British Columbia. Rural, Remote, First Nations and Indigenous COVID-19 Response Framework [Internet]. 2020 [cited 2025 May 18]. Available from: https://www2.gov.bc.ca/assets/gov/public-safety-and-emergency-services/emergency-preparedness-response-recovery/gdx/rural-and-remote-covid-19-response-framework.pdf

32. Australian Digital Health Agency. National Digital Health Strategy 2023-2028. [Internet]. Sydney; 2023 [cited 2025 May 18]. Available from: https://www.digitalhealth.gov.au/sites/default/files/documents/national-digital-health-strategy-2023-2028.pdf

33. National Quality Forum. National Quality Forum. Rural Telehealth and Healthcare System Readiness Measurement Framework – Final Report [Internet]. Washington, DC; 2021 Nov [cited 2025 May 18]. Available from: https://www.qualityforum.org/Publications/2021/11/Rural_Telehealth_and_Healthcare_System_Readiness_Measurement_Framework_-_Final_Report.aspx

34. Butzner M, Cuffee Y. Telehealth Interventions and Outcomes Across Rural Communities in the United States: Narrative Review. J Med Internet Res. 2021 Aug 26;23(8):e29575.

35. Bradford N, Caffery L, Smith A. Telehealth services in rural and remote Australia: a systematic review of models of care and factors influencing success and sustainability. Rural Remote Health. 2016 Oct 17;

36. Thompson AE, Shaw T, Nott S, Wilson A, Saurman E. Patient and carer experiences of hospital-based hybrid virtual medical care: a qualitative study. Medical Journal of Australia. 2024 Dec 9;221(S11).

37. LeBlanc M, Petrie S, Paskaran S, Carson D, Peters P. Patient and provider perspectives on eHealth interventions in Canada and Australia: a scoping review. Rural Remote Health. 2020 Sep 19;

38. Harkey LC, Jung SM, Newton ER, Patterson A. Patient Satisfaction with Telehealth in Rural Settings: A Systematic Review. Int J Telerehabil. 2020 Dec 8;12(2):53–64.

39. Homer R, Biller S, Schumaker B, Johnson PE. Telehealth Usability Among Rural and Low-Income Populations: A Survey of Wyoming Medicaid Members. J Patient Exp. 2024 Jan 23;11.

40. Moecke DP, Holyk T, Beckett M, Chopra S, Petlitsyna P, Girt M, et al. Scoping review of telehealth use by Indigenous populations from Australia, Canada, New Zealand, and the United States. J Telemed Telecare. 2024 Oct 13;30(9):1398–416.

41. Maita KC, Maniaci MJ, Haider CR, Avila FR, Torres-Guzman RA, Borna S, et al. The Impact of Digital Health Solutions on Bridging the Health Care Gap in Rural Areas: A Scoping Review. Perm J. 2024 Sep 16;28(3):130–43.

42. Oliver A, Chandler E, Gillard JA. Impact of Digital Inclusion Initiative to Facilitate Access to Mental Health Services: Service User Interview Study. JMIR Ment Health. 2024 Jul 26;11:e51315.

